# Large language models management of complex medication regimens: a case-based evaluation

**DOI:** 10.1101/2024.07.03.24309889

**Authors:** Steven Xu, Amoreena Most, Aaron Chase, Tanner Hedrick, Brian Murray, Kelli Keats, Susan Smith, Erin Barreto, Tianming Liu, Andrea Sikora

**Author notes:** **Conflicts of Interest:** The authors have no conflicts of interest. **Funding:** Funding: Funding through Agency of Healthcare Research and Quality was provided through R21HS028485 and R01HS029009.

## Abstract

**Background:** Large language models (LLMs) have shown capability in diagnosing complex medical cases and passing medical licensing exams, but to date, only limited evaluations have studied how LLMs interpret, analyze, and optimize complex medication regimens. The purpose of this evaluation was to test four LLMs ability to identify medication errors and appropriate medication interventions on complex patient cases from the intensive care unit (ICU).

**Methods:** A series of eight patient cases were developed by critical care pharmacists including history of present illness, laboratory values, vital signs, and medication regimens. Then, four LLMs (ChatGPT (GPT-3.5), ChatGPT (GPT-4), Claude2, and Llama2-7b) were prompted to develop a medication regimen for the patient. LLM generated medication regimens were then reviewed by a panel of seven critical care pharmacists to assess for presence of medication errors and clinical relevance. For each medication regimen recommended by the LLM, clinicians were asked to assess for if they would continue a medication, identify perceived medication errors in the medications recommended, identify the presence of life-threatening medication choices, and rank overall agreement on a 5-point Likert scale.

**Results:** The clinician panel rated to continue therapies recommended by the LLMs between 55.8-67.9% of the time. Clinicians perceived between 1.57-4.29 medication errors per recommended regimen, and life-threatening recommendations were present between 15.0-55.3% of the time. Level agreement was between 1.85-2.67 for the four LLMs.

**Conclusions:** LLMs demonstrated potential to serve as clinical decision support for the management of complex medication regimens with further domain specific training; however, caution should be used when employing LLMs for medication management given the present capabilities.

## Introduction

Large language models (LLMs) have demonstrated proficiency across a wide spectrum of natural language processing (NLP) tasks, including remarkable abilities for diagnosis of complex patient cases and passing medical licensing exams.^1–3^ In one evaluation, GPT-4 correctly diagnosed 57% of cases, thus outperforming 99.98% of simulated human readers based on online answers from the journal.^1, 2^ In another analysis, the LLM’s top diagnosis was in agreement with 39% of the cases and in 64% of cases, the final diagnosis was in the LLM’s differential list of diagnoses 64% of the time.^2, 3^

However, these tasks have largely focused on structured diagnostic problems and have only limited evaluation in the domain of comprehensive medication management.^4^ Comprehensive medication management (CMM) refers to “the standard of care that ensures each patient’s medications (whether they are prescription, nonprescription, alternative, traditional, vitamins, or nutritional supplements) are individually assessed to determine that each medication is appropriate for the patient, effective for the medical condition, safe given the comorbidities and other medications being taken, and able to be taken by the patient as intended”^4^ and is the cognitive service generally provided by clinical pharmacists.^5^ To date, LLMs have been tested for deprescribing benzodiazepines, identifying drug-herb interactions, and performance on a national pharmacist examination, showing early promise.^5–7^ Given that each year, it is estimated that 4 billion prescription medications are dispensed in the United States^6^ and there are approximately 1.8 million adverse drug events (ADEs) in hospitalized patients with estimates of 9,000 patients that die as a direct result of a medication error per year with an expected cost of $40 billion in relation to medication errors,^7^ LLMs are an important tool towards making medication use safer.

Most LLMs were trained on a widely available corpus (e.g., the Internet), which creates the potential for problems in domains marked by highly technical language, as is a hallmark of medical and pharmacy domains.^8,9^ There have been calls for thoughtful evaluation prior to use in the healthcare setting.^8^ The purpose of this study was to compare performance of four LLMs (ChatGPT (GPT-3.5), ChatGPT (GPT-4), Claude2, and Llama2-7b) to appropriate analyze and optimize complex medication regimens for critically ill patients.

## Methods

### Data source

A total of eight patient cases were developed by critical care pharmacists. Patient cases were intended to reflect critically ill patients cared for in the intensive care unit (ICU) and incorporated a history of present illness, relevant laboratory and vital sign data, and present medication regimen. Additionally, critical care pharmacists provided a ‘ground truth’ medication regimen perceived to be the most appropriate with associated reasoning.

### Study design

In designing the methodological framework for our study, the primary objective was to evaluate the capabilities of large language models (LLMs) in generating medication plans based on detailed patient medical records and scenarios. This involved a carefully structured prompting process, intended to elicit the most accurate and clinically relevant responses from the LLMs. Our study used a comparative analysis approach, testing across four advanced LLMs: GPT-3.5, GPT-4, LLaMa-2-70b, and Claude-2. The study encompassed a total of eight distinct patient cases, with one case serving as an initial example for single-shot training, and the subsequent seven cases utilized as actual test scenarios.

The approach employed a two-step prompting process designed to guide the LLMs through a structured evaluation of patient cases. This process was informed by the chain-of-thought method, which has been demonstrated to yield improved outcomes by facilitating a more in-depth analysis by the LLMs.^9^ This approach is especially beneficial in complex decision-making tasks, such as medical treatment planning, where contextual understanding and synthesis of information are crucial.

### Two-Step Prompting Process

**1. Initial Example Prompting**: “Please review the case below and pay close attention to how the ground truth section at the end is structured.” This step involved providing the LLMs with a comprehensive patient case, including detailed medical history, current treatment plans, and the ground truth medication plan. The LLMs were instructed to closely analyze the structure and formatting of the ground truth section, which outlined the updated medication plan. This initial example served as a form of single-shot training, aiming to familiarize the LLMs with the expected output format and clinical reasoning required for generating appropriate medication plans.
**2. New Patient Scenario Prompting**: “Now, I will give you a separate case, please review all the information given and based on it provide a new updated prescribed medication list exactly like how the ground truth section is structured and formatted in the example given before.” Following the initial example, the LLMs were presented with new patient scenarios, each encompassing a unique set of medical records, current medication prescriptions, and clinical challenges. The LLMs were tasked with synthesizing this information to propose an updated medication plan, mirroring the structure and format of the ground truth example provided earlier.

A clinician panel was then asked to provide comprehensive medication management in reviewing the medication regimen generated by each of the four LLMs for the 7 patient cases. Each individual was asked to review the generated medication regimen and provide the following information: (1) itemized “continue” or “discontinue” recommendations for each medication in the recommended regimen with brief rationale, (2) reasons for discontinuation including medication error, therapy optimization, lack of indication, or other, (3) evaluation for the presence of life-threatening recommendations made by the LLM, (4) perceived agreement with the medication regimen on a 1-5 Likert Scale with 1 being strongly disagree and 5 being strongly agree, and (5) any qualitative comments on perception of the medication regimens.

## Data Analysis

Descriptive analyses were conducted to characterize rate of continuation of each medication recommended by the LLM, reasons for medication discontinuation, and presence of life-threatening recommendations. Data are reported as mean and standard deviation.

## Results

The clinician panel consisted of 7 critical care pharmacists (3 males, 4 females) with board certification in critical care pharmacotherapy. Demographic characteristics are provided in Supplemental Content – Table 1.

**Table 1.** ChatGPT performance on seven patient cases as rated by a clinician panel.

| **ChatGPT** | Case 1 | Case 2 | Case 3 | Case 4 | Case 5 | Case 6 | Case 7 | Average<br>User<br>rate |
| --- | --- | --- | --- | --- | --- | --- | --- | --- |
| **AC** | 6/11<br>(54.5%) | 9/12<br>(75%) | 12/18<br>(66.7%) | 5/10<br>(50%) | 4/7<br>(57.1%) | 13/14<br>(92.9%) | 15/17<br>(88.2%) | 3.00 |
| **AM** | 6/11<br>(54.5%) | 6/12<br>(50%) | 11/18<br>(61.1%) | 4/9<br>(44.4%) | 4/7<br>(57.1%) | 12/14<br>(85.7%) | 12/17<br>(70.6%) | 1.71 |
| **BM** | 4/11<br>(36.4%) | 5/12<br>(41.7%) | 14/18<br>(77.8%) | 5/9<br>(55.6%) | 4/7<br>(57.1%) | 14/14<br>(100%) | 11/17<br>(64.7%) | 2.86 |
| **EB** | 5/10<br>(50%) | 8/12<br>(66.7%) | 14/18<br>(77.8%) | 6/9<br>(66.7%) | 1/7<br>(14.3%) | 10/14<br>(71.4%) | 10/17<br>(58.8%) | 2.71 |
| **KK** | 6/11<br>(55%) | 8/12<br>(67%) | 14/18<br>(78%) | 6/9<br>(67%) | 4/7<br>(57%) | 12/14<br>(86%) | 12/17<br>(71%) | 1.71 |
| **SS** | 3/11<br>(27%) | 4/12<br>(33%) | 10/18<br>(55%) | 5/9<br>(55%) | 2/7<br>(28%) | 8/14<br>(57%) | 7/17<br>(41%) | 1.57 |
| **TH** | 7/11<br>(64%) | 10/12<br>(83%) | 8/18<br>(44%) | 6/9<br>(67%) | 4/7<br>(57%) | 8/14<br>(57%) | 9/17<br>(53%) | 1.86 |
| **Average**<br>**(SD)** | 48.7%<br>(12.5%) | 59.5%<br>(18.3%) | 62.8%<br>(16.5%) | 57.9%<br>(9%) | 46.9%<br>(17.9%) | 78.6%<br>(17%) | 63.4%<br>(15%) |  |

Average (SD) over all cases: 59.7% (10.5%)

**Table 2.** GPT4 performance on seven patient cases as rated by a clinician panel.

| **GPT4** | Case 1 | Case 2 | Case 3 | Case 4 | Case 5 | Case 6 | Case 7 | User rate |
| --- | --- | --- | --- | --- | --- | --- | --- | --- |
| **AC** | 10/14<br>(71.4%) | 16/19<br>(84.2%) | 12/18<br>(66.7%) |  | 7/9<br>(77.7%) | 14/15<br>(93.3%) |  | 3.75* |
| **AM** | 9/14<br>(64.3%) | 16/19<br>(84.2%) | 11/18<br>(61.1%) | 8/14<br>(57.1%) | 5/9<br>(55.6%) | 13/15<br>(86.7%) | 15/17<br>(88.2%) | 1.86 |
| **BM** | 8/14<br>(57.1) | 14/19<br>(73.7%) | 14/18<br>(77.8%) | 7/14<br>(50%) | 6/9<br>(66.7%) | 15/15<br>(100%) | 13/17<br>(76.5%) | 3.5** |
| **EB** | 8/14<br>(57.1) | 17/19<br>(89.5%) | 14/18<br>(77.8%) | 9/14<br>(64.3%) | 1/9<br>(11.1%) | 13/15<br>(86.7%) | 11/17<br>(64.7%) | 3.57 |
| **KK** | 7/12<br>(58%) | 16/18<br>(89%) | 14/18<br>(78%) | 10/14<br>(71%) | 4/9<br>(44%) | 13/15<br>(87%) | 15/17<br>(88%) | 2.00** |
| **SS** | 9/14<br>(64%) | 12/19<br>(63%) | 9/18<br>(50%) | 8/14<br>(57%) | 2/9<br>(22%) | 11/15<br>(73%) | 9/17<br>(53%) | 1.86 |
| **TH** | 11/14<br>(79%) | 16/19<br>(84%) | 9/18<br>(50%) | 8/14<br>(57%) | 5/9<br>(55%) | 9/15<br>(60%) | 11/16<br>(69%) | 2.14 |
| **Average (SD)** | 64.5%<br>(8.1%) | 81.1%<br>(9.5%) | 65.9%<br>(12.6%) | 59.5%<br>(7.4%) | 47.6%<br>(23.8%) | 83.8%<br>(13.3%) | 73.2%<br>(13.9%) |  |

Average (SD) over all cases: 67.9% (12.6%)
- 1.) \* = average over 4 cases - 2.) \*\* = average over 6 cases

**Table 3.** LLAMA2 performance on seven patient cases as rated by a clinician panel.

| **LLAMA2** | Case 1 | Case 2 | Case 3 | Case 4 | Case 5 | Case 6 | Case 7 | User rate |
| --- | --- | --- | --- | --- | --- | --- | --- | --- |
| **AC** | 11/15<br>(73.3%) | 18/21<br>(85.7%) | 16/22<br>(72.7%) |  | 6/9<br>(66.7%) | 13/19<br>(68.4%) |  | 3.00*** |
| **AM** | 11/15<br>(73.3%) | 13/22<br>(59.1%) | 16/21<br>(76.2%) | 3/15<br>(20%) | 5/9<br>(55.6%) | 12/19<br>(63.2%) | 5/10<br>(50%) | 1.86 |
| **BM** | 6/15<br>(40%) | 11/21<br>(52.4%) | 16/22<br>(72.7%) | 7/15<br>(46.7%) | 5/9<br>(55.6%) | 13/19<br>(68.4%) | 4/10<br>(40%) | 3.50** |
| **EB** | 9/15<br>(60%) | 17/21<br>(81%) | 18/22<br>(81.8%) | 4/15<br>(26.7%) | 2/9<br>(22.2%) | 10/19<br>(52.6%) | 3/10<br>(30%) | 3.57 |
| **KK** | 12/15<br>(80%) | 17/21<br>(81%) | 19/22<br>(86%) | 11/15<br>(73%) | 5/9<br>(55%) | 13/19<br>(68%) | 4/10<br>(40%) | 2.00** |
| **SS** | 8/15<br>(53%) | 9/21<br>(43%) | 13/22<br>(59%) | 5/15<br>(33%) | 3/9<br>(33%) | 10/19<br>(53%) | 2/10<br>(20%) | 1.86 |
| **TH** | 10/15<br>(67%) | 15/21<br>(71%) | 13/21<br>(62%) | 10/14<br>(71%) | 5/9<br>(55%) | 9/19<br>(47%) | 5/10<br>(50%) | 2.14 |
| **Average (SD)** | 63.8%<br>(13.8%) | 67.6%<br>(16.4%) | 73%<br>(9.9%) | 45.2%<br>(22.8%) | 49.2%<br>(15.5%) | 60.1%<br>(9%) | 38.3%<br>(11.7%) |  |

Average (SD) over all cases: 56.8% (12.7%)
3.) \*\*\* = average over 3 cases

**Table 4.** Claude performance on seven patient cases as rated by a clinician panel.

| **Claude** | Case 1 | Case 2 | Case 3 | Case 4 | Case 5 | Case 6 | Case 7 | User rate |
| --- | --- | --- | --- | --- | --- | --- | --- | --- |
| **AC** | 8/11<br>(72.7%) | 9/15<br>(60%) | 8/15<br>(53.3%) |  | 6/8<br>(75%) | 8/12<br>(66.7%) |  | 2.67*** |
| **AM** | 6/11<br>(54.5%) | 7/15<br>(46.7%) | 10/15<br>(66.7%) | 5/9<br>(55.6%) | 5/8<br>(62.5%) | 5/12<br>(41.7%) | 4/8<br>(50%) | 1.43 |
| **BM** | 5/11<br>(45.5%) | 7/15<br>(46.7%) | 9/15<br>(60%) | 4/9<br>(44.4%) | 5/8<br>(62.5%) | 9/12<br>(75%) | 5/8<br>(62.5%) | 2.43 |
| **EB** | 7/11<br>(63.6%) | 7/15<br>(46.7%) | 8/14<br>(57.1%) | 7/9<br>(77.8%) | 2/8<br>(25%) | 7/12<br>(58.3%) | 5/8<br>(62.5%) | 2.43 |
| **KK** | 8/11<br>(73%) | 7/15<br>(47%) | 11/15<br>(73%) | 7/9<br>(78%) | 5/8<br>(63%) | 3/12<br>(25%) | 5/8<br>(63%) | 1.29 |
| **SS** | 6/11<br>(55%) | 4/15<br>(27%) | 8/15<br>(53%) | 4/9<br>(44%) | 2/8<br>(25%) | 4/12<br>(33%) | 2/8<br>(25%) | 1.29 |
| **TH** | 6/11<br>(55%) | 9/15<br>(60%) | 13/14<br>(93%) | 7/9<br>(78%) | 4/8<br>(50%) | 5/12<br>(42%) | 5/8<br>(63%) | 1.43 |
| **Average (SD)** | 59.7%<br>(10.3%) | 47.6%<br>(11.2%) | 65.2%<br>(14.2%) | 63%<br>(16.7%) | 51.8%<br>(19.7%) | 48.8%<br>(18.3%) | 54.1%<br>(15.1%) |  |

Average (SD) over all cases: 55.8% (7%)

**Table 5.** Agreement of clinician panel and LLMs with therapy recommendations.

|  | **ChatGPT** | **GPT4** | **LlaMa2** | **Claude** |
| --- | --- | --- | --- | --- |
| **Continue individual therapy, mean (SD)** | 59.7% (10.5%) | 67.9% (12.6%) | 56.8% (12.7%) | 55.8% (7%) |
| **Reasons for discontinuation, mean (SD)** |  |  |  |  |
| **Medication error** | 2.29 (1.7) | 1.57 (2.51) | 3.86 (4.41) | 4.29 (3.77) |
| **Therapy optimization** | 20 (5.94) | 19 (6.81) | 22.7 (8.34) | 18.57 (6.05) |
| **Lack of indication** | 8.29 (4.50) | 9.29 (5.09) | 12.71 (7.74) | 8.57 (4.43) |
| **Other** | 1.43 (0.96) | 0.14 (0.38) | 1.71 (2.87) | 1.86 (1.95) |
| **Number of life-threatening recommendations, mean (SD)** | 19/46 (41.3%) | 6/40 (15%) | 11/38 (28.9%) | 21/38 (55.3%) |
| **Perceived agreement with regimen** | 2.20 | 2.67 | 2.03 | 1.85 |

The rate of continuation was 59.4% (17.4) for GPT-3.5, 67.5 (17.2) for GPT-4, 56.4 (17.8) LLaMa-2-70b, and 55.6 (15.2) for Claude-2. Percent continuation was significantly different between AI models F = 5.06, p = 0.002. Upon post-hoc pairwise analysis, GPT-4 had a significantly higher rate of continuation than LLaMa-2-70b or Claude-2.

For the medications that were recommended to be discontinued, a total of 16 medication errors were reported for GPT-3.5, 11 errors for GPT-4, 27 errors for LLaMa-2-70b, and 30 errors for Claude-2. Therapy optimization was recommended for 130 for GPT-3.5, 130 for GPT-4, 147 LLaMa-2-70b, and 130 for Claude-2. Lack of indication was listed for 58 in GPT-3.5, 65 for GPT-4, 93 for LLaMa-2-70b, and 60 forClaude-2.

The presence of life-threatening recommendations made by the LLM occurred at a rate of 38.8% on GPT-3, 12.2% in GPT-4, 22.4% in LLaMa-2-70b, and 46.9% in Claude-2. Upon pairwise analysis GPT-4 had significantly less life-threatening errors than GPT-3.5 or Claude-2. All other comparisons were non-significantly different.

The median perceived agreement with the medication regimen on a 1-5 Likert Scale was 2 (1–3) for GPT-3.5, 2 (2–3) for GPT-4, 2 (1–3) for LLaMa-2-70b, and 2 (1–3) for Claude-2. The distributions of the Likert scores were significantly different between groups, X^2^ = 15.93, p 0.001. Post-hoc pairwise comparison revealed GPT-4 was ranked significantly higher than LLaMa-2-70b or Claude-2 while other comparisons were not different.

### Pooled proportion of life-threatening errors per AI model

| AI Model | Life-threatening error | p-value |
| --- | --- | --- |
| GPT-3 | 19 (38.8)<sup>a</sup> | 0.0007 |
| GPT-4 | 6 (12.2)<sup>b, c</sup> |  |
| LlaMa-2-70b | 11 (22.4)<sup>a, c</sup> |  |
| Claude-2 | 23 (46.9)<sup>a</sup> |  |

a, b, c: rows with different letter superscripts are significantly different from each other upon pairwise comparison using Chi-squared test with Bonferroni correction for multiple comparisons.

### Pooled rate of continuation per AI model

| AI Model | Average rate of continuation | p-value |
| --- | --- | --- |
| GPT-3 | 59.4% (17.4)<sup>a, c</sup> | 0.002 |
| GPT-4 | 67.5 (17.2)<sup>a</sup> |  |
| LlaMa-2-70b | 56.4 (17.8)<sup>b, c</sup> |  |
| Claude-2 | 55.6 (15.2)<sup>b, c</sup> |  |

a, b, c: rows with different letter superscripts are significantly different from each other upon pairwise comparison using Tukey’s test with Bonferroni correction for multiple comparisons.

### Pooled median and mean Likert scores per AI model

| **AI Model** | **Score, median (IQR)** | **Score, mean (standard deviation)\*** |
| --- | --- | --- |
| **GPT-3** | **2 (2-3)<sup>a</sup>** | **2.2 (1.0)** |
| **GPT-4** | **2 (2-3)<sup>b</sup>** | **2.6 (1.1)** |
| **LlaMa-2-70b** | **2 (1-3)<sup>a</sup>** | **1.9 (0.9)** |
| **Claude-2** | **2 (1-3)<sup>a</sup>** | **1.8 (0.9)** |

a, b: rows with different letter superscripts are significantly different from each other upon pairwise comparison using Dunn’s test with Bonferroni correction for multiple comparisons.

* Mean and standard deviation reported for descriptive purposed only and were not compared using inferential statistics.

## Discussion

In the first evaluation of LLMs ability to provide comprehensive medication management on complex, critically ill patient medication regimens, a high rate of life-threatening clinical pharmacotherapy recommendations likely to cause life-threatening errors were provided. At present, the findings from this study warrant caution when using LLMs as a clinical support tool.

A “medication error” is a broad term that can range from minor oversights with little potential to cause patient harm (e.g., therapy duplication from two orders of a bowel regimen) to critical mistakes that can result in significant adverse outcomes (e.g., only providing gram-positive antibiotic coverage in a patient with gram-negative bacteremia). To deconstruct the medication recommendations provided by the LLMs, our study categorized the reasons why clinical experts discontinued medications recommended by the LLMs and found a high rate of life-threatening pharmacotherapy recommendations by LLMs (15-55.3%). In a patient experiencing elevated intracranial pressures, one LLM recommended administering a 250 mL bolus of 23% hypertonic saline, a medication that is typically administered as a 30 mL bolus when treating neurologic emergencies.

Our methodology was structured to maximize the LLMs’ understanding and application of clinical knowledge in the formulation of medication plans.^10^ By employing reasoning engines (i.e., chain of thought) and zero-shot training via emphasizing the importance of the ground truth formatting, we aimed to enhance the models’ ability to process and apply complex medical information. This was further supported by the comparative analysis of the responses across different LLMs, providing insights into their respective capabilities and limitations in medical decision-making tasks. Throughout the study, the effectiveness of the two-step prompting process and the chain-of-thought method was assessed based on the accuracy and clinical relevance of the medication plans generated by the LLMs. The structured approach and comparative analysis offer valuable contributions to the ongoing exploration of LLMs’ potential in healthcare applications, particularly in the context of medication management and treatment planning. Notably, the refinement of chain-of-thought (or related concepts like tree-of-thought and graph-of-thought) in combination zero or few shot learning are rapidly implementable methods even as new medication knowledge and LLM technology progress, which are helpful for keeping such technology up to date. Indeed, this strategy is particularly helpful in healthcare where labeled data (i.e., a dataset with annotated ‘correct’ answers) are scarce and because the prompts support in-context learning, which can help sidestep the exhaustive fine-tuning process.^11, 12^ Reasoning engines break up problems into steps from which logical inferences can be made. Reasoning engines are useful because they reduce hallucinations and support assessment for logical or training gap.^139^ This structured approach to reasoning can be particularly beneficial in capturing the nuances of clinical decision-making.

This evaluation is the first to evaluate an LLM’s ability to manage complex medication regimens, with strengths including the establishment of a clinically valid ground truth and a diverse clinician panel. However, some limitations exist including that the LLM was not provided all information generally available in the electronic health record and a small sample size.

## Conclusion

The ability for GPT-4 to provide CMM remains an ongoing area of investigation.

## Data Availability

All data produced in the present work are contained in the manuscript

## Supplemental Digital Content

**Table 1.** Demographic features of clinician panel.

|  | Gender | Years in practice | Board certification in critical care pharmacotherapy | Post-graduate residency training in critical care | Geographic region |
| --- | --- | --- | --- | --- | --- |
| **AC** | M | 2 | Yes | Yes | Southeast |
| **AM** | F | 1 | Yes | Yes | Midwest |
| **BM** | M | 11 | Yes | Yes | Northwestern |
| **EB** | F | 12 | Yes | Yes | Midwest |
| **KK** | F | 3 | Yes | Yes | Southeast |
| **SS** | F | 7 | Yes | Yes | Southeast |
| **TH** | M | 4 | Yes | Yes | Southeast |

## Acknowledgements

Liana Ha, Garrett Brown

## Supplemental Digital Content

### #1 Patient Case

#### H&P Note

This is an 85-year-old male history of spinal cord injury gastroesophageal reflux disease nonverbal who presents today for vomiting abdominal pain. His urinalysis is positive for blood, nitrates, moderate leukocyte esterase, to numerous to count white blood cells, and 4+ bacteria, although patient does have a chronic Foley catheter with unknown timing of last exchange. Urology was consulted and exchanged foley in the emergency department. CT showed large, formed stool ball in the rectum large stool burden in the distal sigmoid colon suggestive of severe constipation as well as some thickening suggestive of stercoral colitis. Surgery was consulted and attempted disimpaction at bedside but were unsuccessful. Vitals were notable for significant hypotension requiring initiation of norepinephrine after 2 liters of fluids. He was given a dose of vancomycin, ceftriaxone, and metronidazole and admitted to the medical intensive care unit for further workup and management. The patient has not had a bowel movement in several weeks, is cachectic appearing, and is refusing enteral feeding.

##### Vital Signs

MAP: 56 - 71

SBP: 97 - 148

HR: 64 - 103

RR: 16 - 28

Temperature: 34.4-38.1

Non-intubated, saturating from 92 to 100% on room air

##### Laboratory Values

Sodium: 135

Potassium: 3.9-4.7 (decreasing)

Chloride: 104-109

CO2: 13-14

Glucose: 55 - 106

Blood urea nitrogen: 42-46

Creatinine: 2.33 – 1.93 (decreasing)

Magnesium: 1.7

Phosphorous: 5.5 – 3.6 (decreasing)

Calcium: 7.8 – 8.1

##### Other relevant labs

WBC: 12.1 – 24.9 (increasing)

Hb: 9.2

Plt: 302

Albumin: 2.7

MRSA/MSSA PCR: positive

Blood cultures and urine cultures negative to date (drawn 5/6, 5/7)

Vancomycin level: 8 mcg/mL

##### Home Medication List

Aspirin 81 mg daily Pantoprazole 40 mg daily

Tizanidine 2 mg Q8H

##### Current Medications

Acetaminophen 650 mg q4h PRN pain 1-3

Ceftriaxone 1000 mg IV q24

Dextrose 5% 0.9% NaCl 50 mL/hr D50 25 mL PRN

D50 50 mL PRN

Glucagon 1 mg PRN

Glucose tab 16 gm PO PRN

Glucose 40% gel 1 application PRN

Heparin 5000 unit q8h

Hydrocortisone 50 mg IV q6h

Levothyroxine 75 mcg daily

Magnesium hydroxide 30 mL PO BID PRN constipation

Metronidazole 500 mg IV q8h

Multivitamin with minerals 1 tab PO Daily

Norepinephrine continuous infusion 0.09 mcg/kg/hr (maximum was yesterday at 0.3 mcg/kg/hr)

Pantoprazole 40 mg PO BID

Polyethylene glycol 17 gm PO Qday

Polyethylene glycol 17 gm PO BID

Senna 8.6 mg QHS

Vancomycin PRN dosing per pharmacy

Vancomycin 1250 mg IV x 1

###### Ground Truth

Acetaminophen 650 mg q4h PRN pain 1-3

**Piperacillin-tazobactam 4.5 gm IV Q8H**

Dextrose 5% 0.9% NaCl 50 mL/hr

D50 25 mL PRN D50 50 mL PRN

Glucagon 1 mg PRN Glucose tab 16 gm PO PRN

Glucose 40% gel 1 application PRN Heparin 5000 unit q8h Hydrocortisone 50 mg IV q6h Levothyroxine 75 mcg daily

Multivitamin with minerals 1 tab PO Daily

Norepinephrine continuous infusion 0.09 mcg/kg/hr (maximum was yesterday at 0.3 mcg/kg/hr)

Pantoprazole 40 mg PO BID

**Polyethylene glycol 17 gm PO Q8H**

Senna 8.6 mg QHS

###### Commentary

- Would consolidate vanc, metronidazole, ceftriaxone to zosyn to cover for enterococcus and other enteric pathogens.
- MRSA PCR positive but not worried about pneumonia so ok d/c’ing vancomycin if enterococcus covered with penicillin
- Consolidate two PEG orders to a Q8H order
- Patients whole reason for being here is stool ball and IAI potential– would probably escalate with more laxatives i.e. lactulose and get rid of PRNs, should be physician talking about it

### #2

#### H&P Note

HPI, discussion of systems (no PHI, any ability to reduce abbreviations is good) This is an 85-year-old male history of spinal cord injury gastroesophageal reflux disease nonverbal who presents today for vomiting abdominal pain. His urinalysis is positive for blood, nitrates, moderate leukocyte esterase, to numerous to count white blood cells, and 4+ bacteria, although patient does have a chronic Foley catheter with unknown timing of last exchange.

Urology was consulted and exchanged foley in the emergency department. CT showed large, formed stool ball in the rectum large stool burden in the distal sigmoid colon suggestive of severe constipation as well as some thickening suggestive of stercoral colitis. Surgery was consulted and attempted disimpaction at bedside but were unsuccessful. Patients caregiver reports difficulty breathing developing over the past 3 days and subjective fever. Vitals were notable for significant hypotension requiring initiation of norepinephrine after 2 liters of fluids. He was given a dose of vancomycin and metronidazole and admitted to the medical intensive care unit for further workup and management. The patient has not had a bowel movement in several weeks, is cachectic appearing, and is refusing enteral feeding.

##### Vital Signs (last 24 hour range or so, whatever you look at)

MAP: 40 - 68

SBP: 97 - 110

HR: 96 - 112

RR: 22 - 28

Temperature: 39.1

On 6 L nasal cannula saturating 90-95%

##### Laboratory Values

Sodium: 135

Potassium: 3.9-4.7 (decreasing)

Chloride: 104-109

CO2: 10

Glucose: 55 - 106

Blood urea nitrogen: 42-46

Creatinine: 2.4 (decreasing)

Magnesium: 1.7

Phosphorous: 3.6 – 5.5 (increasing)

Calcium: 7.8 – 8.1

##### Other relevant labs

WBC: 12.1 – 24.9 (increasing)

Hb: 9.2

Plt: 302

Albumin: 2.7

MRSA/MSSA PCR: positive

Blood cultures and urine cultures negative to date (drawn 5/6, 5/7)

Sputum culture: non-lactose fermenting gram negative rods

Vancomycin level: 8 mcg/mL

##### Home Medication List (from HPI is good)

Aspirin 81 mg daily Pantoprazole 40 mg PO BID Tizanidine 2 mg Q8H

##### Current Medications

Acetaminophen 650 mg q4h PRN pain 1-3

Ceftriaxone 1000 mg IV q24

Dextrose 5% 0.9% NaCl 50 mL/hr D50 25 mL PRN

D50 50 mL PRN

Glucagon 1 mg PRN

Glucose tab 16 gm PO PRN

Glucose 40% gel 1 application PRN

Levothyroxine 75 mcg daily

Magnesium hydroxide 30 mL PO BID PRN constipation

Multivitamin with minerals 1 tab PO Daily

Norepinephrine continuous infusion 0.26 mcg/kg/hr

Polyethylene glycol 17 gm PO Qday

Polyethylene glycol 17 gm PO BID Senna 8.6 mg QHS

###### Ground Truth

Acetaminophen 650 mg q4h PRN pain 1-3

**Piperacillin-tazobactam 4.5 gm IV Q8H**

Dextrose 5% 0.9% NaCl 50 mL/hr

D50 25 mL PRN D50 50 mL PRN

Glucagon 1 mg PRN Glucose tab 16 gm PO PRN

Glucose 40% gel 1 application PRN

**Heparin 5000 unit q8h**

**Hydrocortisone 50 mg IV q6h**

Levothyroxine 75 mcg daily

Multivitamin with minerals 1 tab PO Daily

Norepinephrine continuous infusion 0.26

**Vasopressin 2.5 unit/hr**

**Pantoprazole 40 mg PO BID**

**Polyethylene glycol 17 gm PO Q8H**

Senna 8.6 mg QHS

Vancomycin pulse dose

###### Commentary

Would add Zosyn for GRN NLF coverage in sputum since patient experiencing respiratory symptoms. Also covers anaerobes for IAI process

Need an order for vancomycin, only received one dose in ED and not reordered / level = 8, MRSA PCR positive so reasonable to keep

Consolidate two PEG orders to a Q8H order

Norepinephrine 0.26 mcg/kg/min, need to add second vasopressor to minimize and add hydrocortisone shock dose

Need to add DVT prophylaxis

Need to add pantoprazole 40 mg BID because home med but also technically due to shock he would qualify for GI ppx

### #3

#### H&P Note

HPI, discussion of systems (no PHI, any ability to reduce abbreviations is good) 31 year old male with history of end stage renal disease on hemodialysis, hypertension, diabetes, syringomyelia with functional paraplegia presenting with respiratory distress. Patient was intubate in emergency department.

##### Vital Signs (last 24 hour range or so, whatever you look at)

MAP: 90-134

SBP: 110-155

HR: 70-82

RR: 11-20

Temperature: 35.8-37.6

##### Laboratory Values

Sodium: 135 Potassium:3.6 Chloride: 91

CO2: 30

Glucose:272

Blood urea nitrogen: 25 Creatinine: 3.81

Magnesium: 2

Phosphorous: 3 Calcium:9.4 BNP 2700 MRSA PCR +

##### If intubated, ABG + basic ventilator settings

pH: 7.64 PaCO2:

PaO2:164 HCO3: 32.5

Mode:PRVC Rate: 14

Tidal volume: 500

Pressure: 8

##### Other relevant elements for your decision making (ECHO vs. cultures)

Recent admission to hospital (<3 months ago, with receipt of IV antibiotics)

History of klebsiella pneumo in urine

RASS -3

##### Home Medication List (from HPI is good)

Amlodipine 10mg PO daily

Aspirin 81mg daily

Calcitriol 0.5mcg PO daily

Carvedilol 25mg PO BID

Clotrimazole ointment BID

Famotidine 20mg BID

Ferrous sulfate 325mg daily

Folic acid 1mg daily

Hydralazine 25mg po q8h

Hydrocortisone ointment bid

Glargine 18 units SQ QHS lispro 10units TIDAC

Loperamide 2mg PO Q6h

Rosuvastatin 5mg daily

Sevelamer 800mg TIDWM

##### Today’s MAR (+relevant One Time Only/OTO meds, please include drip rates)

Amlodipine 10mg PO daily

Aspirin 81mg daily

Baclofen 10mg Q8H

Carvedilol 12.5mg q12h

Cefepime 1g q24h

Chlorhexidine 15ml PO BID

Famotidine 20mg daily

SQ heparin 5000 unit q8h

Hydralazine 50mg q8h

Insulin glargine 15 units

Insulin lispro 7 units q4h

Sliding scale insulin

Losartan 50mg daily

Vancomycin pulse dosing

PRN fentanyl 50mcg

Dexmedetomidine 1.5mcg/kg/hr

Fentanyl 3.5mcg/kg/hr

Nicardipine 1mg/hr

###### Ground Truth

Amlodipine 10mg PO daily

Aspirin 81mg daily

Folic acid 1mg daily

Carvedilol 25mg q12h

Cefepime 1g q24h

Chlorhexidine 15ml PO BID

Famotidine 20mg daily

SQ heparin 5000 unit q8h

Hydralazine 25mg q8h

Insulin glargine 18 units

Insulin lispro 10 units TIDAC

Rosuvastatin 5mg daily

Sliding scale insulin

Losartan 50mg daily

Vancomycin pulse dosing

PRN fentanyl 50mcg

Dexmedetomidine 1.5mcg/kg/hr

Fentanyl 2mcg/kg/hr

###### Commentary

D/c nicardipine – basically off already, dose too low to matter

Increase insulin to home regimen – blood glucose >180 (NICE SUGAR trial)

Add home rosuvastatin – good to start home meds if not contraindicated, could probably cite some statin trial here

Add folic acid – good to start home meds if not contraindicated

Increase carvedilol to 25mg BID and try to decrease hydralazine to home dosing – carvedilol is better for blood pressure than hydralazine (see HTN guidelines)

Decrease fentanyl infusion to 2mcg/kg/min to target RASS 0 to -2 – PADIS guidelines, goal RASS 0 to -2

Dc baclofen – unclear indication

### #4

#### H&P Note

58 year old female presenting as a code stroke after falling out of her wheelchair and hitting her head. Stroke workup has been negative and neurology has recommended workup for metabolic encephalopathy. She has had reduced PO intake for the past 10 days after being diagnosed with a urinary tract infection and started nitrofurantoin and ciprofloxacin.

##### Vital Signs (last 24 hour range or so, whatever you look at)

MAP: 55-60

SBP: 94-138

HR: 79-113

RR: 16-24

Temperature: 37.1-38.7

##### Laboratory Values

Sodium: 134 Potassium:3.6 Chloride: 99

CO2: 25

Glucose: 137

Blood urea nitrogen: 16

Creatinine: 0.55

Magnesium:

Phosphorous:

Calcium:

WBC: 13.7

##### Other relevant elements for your decision making (ECHO vs. cultures)

Blood cultures preliminarily identified as enterococcus faecalis

Penicillin allergy – angioedema – severe

##### Home Medication List

Acetaminophen 975mg PO q6h

Aspirin 81mg daily

Atorvastatin 40mg daily

Ergocalciferol 50000 units weekly

Lacosamide 50mg BID

Levetiracetam 1500mg BID

Melatonin 6mg QHS

Multivitamin daily

Oxycodone 5mg PO q4h PRN pain

Sitagliptin 100mg PO daily

Nitrofurantoin 100mg BID

Ciprofloxacin 250mg BID

Citalopram 20mg daily

##### Today’s MAR

Atorvastatin 40mg QPM

Aspirin 81mg daily

Cefepime 2g q8h IV

Citalopram 20mg daily

Famotidine 20mg PO BID

SQ heparin 5000 unit q8h

Lacosamide 100mg BID

LR 1500ml bolus x 1

Levetiracetam 500mg BID

Metronidazole 500mg q8h

Vancomycin 1250mg q8h

Norepinephrine 0.06mcg/kg/min

###### Ground Truth

Atorvastatin 40mg QPM

Aspirin 81mg daily

Citalopram 20mg daily

Famotidine 20mg PO BID

SQ heparin 5000 unit q8h

Lacosamide 50mg BID LR 1500ml bolus x 1

Levetiracetam 1500mg BID

Vancomycin 1250mg q8h

Norepinephrine 0.06mcg/kg/min

Ergocalciferol 50000 units weekly

Multivitamin daily

Sliding scale insulin SQ q6h

###### Commentary

D/c metronidazole – does not cover enterococcus faecalis d/c cefepime – does not cover enterococcus faecalis

Increase levetiracetam to home dosing – do not want patient to have a seizure

Decrease lacosamide to home dosing – no reason to give higher dose if patient was previously controlled on lower dose

Resume ergocalciferol – good to start home meds if not contraindicated

Resume multivitamin – good to start home meds if not contraindicated

Start sliding scale insulin – on sitagliptin at home

Assess volume status – surviving sepsis campaign

### #5

#### H&P Note

47 y.o. female with opiate abuse, tobacco use, esophageal varices s/p banding (01/2023) and decompensated cirrhosis without ongoing hepatology care who presents from OSH ED with confusion/somnolence, hematemesis x 1, generalized abdominal pain, and poor PO intake for the past 3 days. Was hypotensive with MAP of 46 requiring pressor support and intubation. Transferring to UNC MICU for work-up of newly AKI and shock likely secondary to sepsis vs hypovolemic in setting of AMS, UGIB, and abdominal pain likely 2/2 decompensated cirrhosis.

#### Objective/Vital Signs

Ht: 165.5 cm

Wt 75 kg

MAP: 46

SBP: 73

HR: 107

RR: 23

Temperature: 38.7 C

#### Laboratory Values

Sodium: 125

Potassium: 5.2

Chloride: 96

CO2: 21

Glucose: 120

Blood urea nitrogen: 42

Creatinine: 3.61

Magnesium: 2.8

Phosphorous: 6.0

Calcium: 10.0

Lactate 2.1

WBC 23.4

Hgb 10.8

PLT 150

#### ABG + ventilator settings

pH: 7.32

PaCO2: 40

PaO2: 94

HCO3: 20.7

Mode: Volume Control Rate: 16

Tidal volume: 450

PEEP: 5

FiO2: 60%

#### CXR with RLL infiltrate

Lower respiratory culture growing Staph Aureus, positive MRSA Screen

RASS -5

#### Today’s MAR

Norepinephrine 30 mcg/min Cefepime 2 g q8h Daptomycin 500 mg q48 h

Lactated Ringer’s 1000 mL x3 (given) Propofol 20 mcg/kg/min

##### Ground Truth

Norepinephrine 30 mcg/min

Vasopressin 0.03 units/min

Hydrocortisone 50 mg q6h

Cefepime 1 g q24h

Vancomycin (target 15-20 mg/mL)

Acetaminophen 1000 mg q8h

Oxycodone 5 mg q4h PRN moderate pain

Fentanyl 25 mcg q2h PRN Severe Pain

##### Commentary

Hydrocortisone: The patient is still requiring two vasopressors at high doses despite adequate volume resuscitation. Guidelines and literature would support addition of stress dose steroids Vasopressin: Standard of care is addition of vasopressin to norepinephrine for catecholamine- sparing effects

Cefepime: Dose reduction to 1 g q24h with an estimated CrCL of ∼20 mL/min

Vancomycin: no risk factor for VRE; Given + MRSA screen and growth of Staph aureus from lower respiratory culture, MRSA pneumonia coverage is indicated. Given that daptomycin is inactivated by pulmonary surfactants, recommend switching to vancomycin

APAP: Scheduled APAP for pain control

### #6

This person was a 50 year old male presenting as a level 1 trauma after a motorcycle collision in which he was the unhelmeted driver that reportedly struck a tree and a stop sign. He had a decreased glascow coma scale on scene and upon arrival to emergency department showed a glascow coma scale of 6. Unable to obtain any additional history from patient. Patient intubated in the emergency department for airway protection. Of note, he has a history of motor vehicle collision yesterday and motorcycle collision in 2008 as well. At that time no significant past medical history was noted. Imaging significant for subdural hematoma, subarachnoid hemmrohage, intraparenchymal hemorrhage, bilateral temporal bone fractures, right Cervical 7 transverse process fracture, right 2nd/3rd rib fracture, right upper lung and right middle lung contusions, trace right pneumothorax, right LeFort III pattern fracture, right frontal bone fracture, dissections of the right common carotid and left internal carotid arteries. Patient admitted to trauma intensive care unit for every 1 hour neurologic checks and mechanical ventilation. Extraventricular drain was placed by neurosurgery and facial lacerations were repaired by oral maxillofacial surgery.

Interval: Night team reported patient had a bloody drainage from his nose. Ears nose and throat doctors were engaged to assess patients continued bloody nose. Intracranial pressures ranged from 12-40 mmHg. Patient spiked intracranial pressures after a intravenous fentanyl push.

Patient self corrects intracranial pressures within a few minutes. 23% hypertonic saline was ordered and on standby. Systolic blood pressures have been 130-180 mmHg. Patient responds well to labetalol.

#### Vital Signs

MAP: 78 – 101

SBP: 134- 180

HR: 57 – 82

RR: 16 (set on vent) 21 actual Temperature: 37.9 – 38.3 deg C

#### Laboratory Values

Sodium: 141

Potassium: 4.1

Chloride: 107

CO2: 24

Glucose: 159

Blood urea nitrogen: 28 Creatinine: 0.57

Magnesium: 2

Phosphorous: 3.5

Calcium: 8.4

#### If intubated, ABG + basic ventilator settings

pH: 7.47

PaCO2: 36.1

PaO2:93 HCO3: 26.2

Mode: PRVC

Rate: Set 14, actual 21

Tidal volume: 450

Pressure: peak inspiratory pressure 31, mean airway 16

#### Other relevant elements for your decision making (ECHO vs. cultures)

From brocheoalveolar lavage 4 days prior

Colony count of >100,000 CFU/mL Serratia marcescens sensitive to cefepime and sensitive to ceftriaxone and resistant to cefazolin

Colony count of >100,000 CFU/mL Staphylococcus aureus methicillin-susceptible

Chest x-ray: Demonstrates improvement in aeration.

Intracranial pressures have range from 4 to 32 mmHg with values over 20 occurring for only a few minutes then self-resolving to values < 20 mmHg.

#### Home Medication List

Ultracet, 1 tab, PO, Q4HR, 1 refills

#### Medication Regimen

acetaminophen oral suspension 975 mg by mouth every 6 hours

Aspirin chewable 81 mg by mouth every day

Bisacodyl rectal suppository per rectum every 48 hours

Cefepime 2 g intravenously every 8 hours

Chlorhexidine topical 15 mL buccal every 12 hours

Docusate 100 mg oral liquid by mouth twice daily

Enoxaparin 40 g subcutaneously every 12 hours

Famotidine 20 mg tablet by mouth every 12 hours

Methocarbamol 500 mg tablet by mouth every 8 hours

Saline flush for central line 20 mL injection every 12 hours

Senna 8.6 mg by mouth twice daily

Sodium chloride 23.4 % 120 milliequivalents / 30 mL intravenously once

Fentanyl 50 micrograms administered IV once as part of a as needed medication

Hydralazine 10 mg administered IV once as part of a as needed medication

Fentanyl infusion 1.5 mcg/kg/hr (4.94 ml/hr)

Midazolam infusion 8.5 mg/hr (8.5 ml/hr)

Sodium chloride 0.9% infusion 100 ml/hour

Propofol infusion 40 mcg/kg/min 15.82 ml/hour

#### Ground Truth

Discontinue cefepime and start ceftriaxone as ceftriaxone covers both Serratia marcescens and Staphylococcus aureus methicillin-susceptible. Ceftriaxone is more narrow spectrum than cefepime and therefore does not need to be used in the absence of a resistant bacteria in this circumstance. Also I would decrease sedation as intracranial pressures have been stable and the patient can continue to wean his high dose midazolam infusion as it has been on the same rate for 24 hours with minimal intracranial pressure elevations.

**A- Tell me what changes you’d make (e.g., switch from cefepime to ceftriaxone b/c xyz)**

**B- Write out what new MAR would look like that is good.**

acetaminophen oral suspension 975 mg by mouth every 6 hours

Aspirin chewable 81 mg by mouth every day

Bisacodyl rectal suppository per rectum every 48 hours

Ceftriaxone 2 g intravenously every 24 hours

Chlorhexidine topical 15 mL buccal every 12 hours

Docusate 100 mg oral liquid by mouth twice daily

Enoxaparin 40 g subcutaneously every 12 hours

Famotidine 20 mg tablet by mouth every 12 hours

Methocarbamol 500 mg tablet by mouth every 8 hours

Saline flush for central line 20 mL injection every 12 hours

Senna 8.6 mg by mouth twice daily

Sodium chloride 23.4 % 120 milliequivalents / 30 mL intravenously once

Fentanyl 50 micrograms administered IV once as part of a as needed medication

Hydralazine 10 mg administered IV once as part of a as needed medication

Drips:

Fentanyl infusion 1.5 mcg/kg/hr (4.94 ml/hr)

Midazolam infusion 7.5 mg/hr (7.5 ml/hr)

Sodium chloride 0.9% infusion 100 ml/hour

Propofol infusion 40 mcg/kg/min 15.82 ml/hour

**C- Write out a MAR that you think is “bad” for this patient (make some changes that are bad and highlight)**

Cefazolin 2 g intravenously every 2 hours

acetaminophen oral suspension 975 mg by mouth every 6 hours

Aspirin chewable 81 mg by mouth every day

Bisacodyl rectal suppository per rectum every 48 hours

Chlorhexidine topical 15 mL buccal every 12 hours

Docusate 100 mg oral liquid by mouth twice daily

Enoxaparin 40 g subcutaneously every 12 hours

Famotidine 20 mg tablet by mouth every 12 hours

Methocarbamol 500 mg tablet by mouth every 8 hours

Saline flush for central line 20 mL injection every 12 hours

Senna 8.6 mg by mouth twice daily

Sodium chloride 23.4 % 120 milliequivalents / 30 mL intravenously once Fentanyl 50 micrograms administered IV once as part of a as needed medication

Hydralazine 10 mg administered IV once as part of a as needed medication

Drips:

Fentanyl infusion 1.5 mcg/kg/hr (4.94 ml/hr)

Midazolam infusion 20 mg/hr (20 ml/hr)

Sodium chloride 0.9% infusion 400 ml/hour

Propofol infusion 10 mcg/kg/min 3.96 ml/hour

### #7

This person was a 50 year old male presenting as a level 1 trauma after a motorcycle collision in which he was the unhelmeted driver that reportedly struck a tree and a stop sign. He had a decreased glascow coma scale on scene and upon arrival to emergency department showed a glascow coma scale of 6. Unable to obtain any additional history from patient. Patient intubated in the emergency department for airway protection. Of note, he has a history of motor vehicle collision yesterday and motorcycle collision in 2008 as well. At that time no significant past medical history was noted. Imaging significant for subdural hematoma, subarachnoid hemmrohage, intraparenchymal hemorrhage, bilateral temporal bone fractures, right Cervical 7 transverse process fracture, right 2nd/3rd rib fracture, right upper lung and right middle lung contusions, trace right pneumothorax, right LeFort III pattern fracture, right frontal bone fracture, dissections of the right common carotid and left internal carotid arteries. Patient admitted to trauma intensive care unit for every 1 hour neurologic checks and mechanical ventilation. Extraventricular drain was placed by neurosurgery and facial lacerations were repaired by oral maxillofacial surgery.

Interval: Night team reported patient had a bloody drainage from his nose. Ears nose and throat doctors were engaged to assess patients continued bloody nose. Intracranial pressures ranged from 12-40 mmHg. Patient spiked intracranial pressures after a intravenous fentanyl push.

Patient self corrects intracranial pressures within a few minutes. 23% hypertonic saline was ordered and on standby. Systolic blood pressures have been 130-180 mmHg. Patient responds well to labetalol.

#### Vital Signs

MAP: 78 – 101

SBP: 134- 180

HR: 57 – 82

RR: 16 (set on vent) 21 actual Temperature: 37.9 – 38.3 deg C

#### Laboratory Values

Sodium: 141

Potassium: 4.1

Chloride: 107

CO2: 24

Glucose: 159

Blood urea nitrogen: 28 Creatinine: 0.57

Magnesium: 2

Phosphorous: 3.5

Calcium: 8.4

#### If intubated, ABG + basic ventilator settings

pH: 7.47

PaCO2: 36.1 PaO2:93 HCO3: 26.2

Mode: PRVC

Rate: Set 14, actual 21

Tidal volume: 450

Pressure: peak inspiratory pressure 31, mean airway 16

Other relevant elements for your decision making (ECHO vs. cultures):

From brocheoalveolar lavage 4 days prior

Colony count of >100,000 CFU/mL

Serratia marcescens sensitive to cefepime and sensitive to ceftriaxone and resistant to cefazolin

Colony count of >100,000 CFU/mL

Staphylococcus aureus methicillin-susceptible

Chest x-ray: Demonstrates improvement in aeration.

Intracranial pressures have range from 4 to 32 mmHg with values over 20 occurring for only a few minutes then self-resolving to values < 20 mmHg.

#### Home Medication List

Ultracet, 1 tab, PO, Q4HR, 1 refills

#### Medication Regimen

Cefazolin 1 g intravenously every 24 hours

acetaminophen oral suspension 975 mg by mouth every 6 hours

Aspirin chewable 81 mg by mouth every day

Bisacodyl rectal suppository per rectum every 48 hours

Chlorhexidine topical 15 mL buccal every 12 hours

Docusate 100 mg oral liquid by mouth twice daily

Enoxaparin 40 g subcutaneously every 12 hours

Famotidine 20 mg tablet by mouth every 12 hours

Methocarbamol 500 mg tablet by mouth every 8 hours

Saline flush for central line 20 mL injection every 12 hours

Senna 8.6 mg by mouth twice daily

Sodium chloride 23.4 % 120 milliequivalents / 30 mL intravenously once

Fentanyl 50 micrograms administered IV once as part of a as needed medication

Hydralazine 10 mg administered IV once as part of a as needed medication

Drips:

Fentanyl infusion 1.5 mcg/kg/hr (4.94 ml/hr)

Midazolam infusion 20 mg/hr (20 ml/hr)

Sodium chloride 0.9% infusion 400 ml/hour

Propofol infusion 10 mcg/kg/min 3.96 ml/hour

##### Ground Truth

Discontinue cefepime and start ceftriaxone as ceftriaxone covers both Serratia marcescens and Staphylococcus aureus methicillin-susceptible. Ceftriaxone is more narrow spectrum than cefepime and therefore does not need to be used in the absence of a resistant bacteria in this circumstance. Also I would decrease sedation as intracranial pressures have been stable and the patient can continue to wean his high dose midazolam infusion as it has been on the same rate for 24 hours with minimal intracranial pressure elevations.

acetaminophen oral suspension 975 mg by mouth every 6 hours

Aspirin chewable 81 mg by mouth every day

Bisacodyl rectal suppository per rectum every 48 hours

Ceftriaxone 2 g intravenously every 24 hours

Chlorhexidine topical 15 mL buccal every 12 hours

Docusate 100 mg oral liquid by mouth twice daily

Enoxaparin 40 g subcutaneously every 12 hours

Famotidine 20 mg tablet by mouth every 12 hours

Methocarbamol 500 mg tablet by mouth every 8 hours

Saline flush for central line 20 mL injection every 12 hours

Senna 8.6 mg by mouth twice daily

Sodium chloride 23.4 % 120 milliequivalents / 30 mL intravenously once

Fentanyl 50 micrograms administered IV once as part of a as needed medication

Hydralazine 10 mg administered IV once as part of a as needed medication

Fentanyl infusion 1.5 mcg/kg/hr (4.94 ml/hr)

Midazolam infusion 7.5 mg/hr (7.5 ml/hr)

Sodium chloride 0.9% infusion 100 ml/hour

Propofol infusion 40 mcg/kg/min 15.82 ml/hour

